# P-TAU205 IS A BIOMARKER LINKED TO TAU-PET ABNORMALITY: A CROSS-SECTIONAL AND LONGITUDINAL STUDY

**DOI:** 10.1101/2025.10.21.25338437

**Authors:** Juan Lantero-Rodriguez, Shorena Janelidze, Sebastian Palmqvist, Luisa Sophie Braun-Wohlfahrt, Jakub Vavra, Divya Bali, Anna Orduña Dolado, Niklas Mattsson-Carlgren, Erik Stomrud, Henrik Zetterberg, Kaj Blennow, Oskar Hansson, Laia Montoliu-Gaya, Gemma Salvadó

## Abstract

Current fluid biomarkers for Alzheimer’s disease (AD) track amyloid-β (Aβ) pathology more strongly than tau, even though clinical and cognitive decline relate more closely to tau. We evaluated cerebrospinal fluid (CSF) phosphorylated tau at epitope 205 (p-tau205), measured using immunoassays, as a biomarker of tau aggregation. A total of 2,069 samples from the BioFINDER-2 (n=1,364) and BioFINDER-1 (n=705) cohorts spanning the full AD continuum were analyzed to assess cross-sectional and longitudinal associations with imaging and clinical measures. CSF p-tau205 levels were elevated in both biologically and clinically advanced disease stages. In Aβ-positive individuals, cross-sectional p-tau205 correlated with Aβ-PET (R²=0.26), tau-PET (R²=0.29), cortical atrophy (R²=0.14) and cognition (MMSE, R²=0.14). Baseline p-tau205 predicted subsequent Aβ accumulation (R²=0.44) and tau-PET uptake (R²=0.33), and increased more steeply over time in Aβ-positive than Aβ-negative participants (β[95%CI]=0.16[0.12–0.21], p<0.001). Longitudinal p-tau205 change related to cortical thinning (R²=0.32) and cognitive decline (R²≥0.41). Incorporating Aβ42/40, p-tau217 and p-tau205 into a CSF-based staging model, the final p-tau205-positive stage showed the strongest cortical atrophy, cognitive impairment, and risk of incident dementia (HR=6.40[4.28–9.59]). These findings support CSF p-tau205 as a valuable marker for biological staging and progression monitoring in AD.

## INTRODUCTION

Alzheimer’s disease (AD) is a slow progressing neurodegenerative disease characterized by a long preclinical phase [1] during which, the two main pathologies, amyloid-β (Aβ) plaques and neurofibrillary tau tangles (NFTs), progressively accumulate eventually causing neuronal dysfunction, neurodegeneration, and cognitive decline [2–4]. In living patients, clinical diagnosis increasingly relies on fluid and imaging biomarkers to assist the evaluation of clinical symptoms [5]. These biomarkers, which can identify and track underlying AD pathology even prior to symptom onset [6–10], have clinical applications that extend beyond diagnosis, including patient management and monitoring, and disease prognosis [9–12]. Moreover, with the recent approval of disease modifying therapies, AD biomarkers are expected to perform a critical role in both clinical trial recruitment and patient selection for disease-modifying treatments [5].

One of the most recent and impactful additions to the AD biomarker portfolio is biofluid measurements of phosphorylated tau at threonine 205 (p-tau205) [13–18]. The concept of measuring this specific phosphorylation originates from post-mortem Braak staging, which is performed by immunostaining using AT8 antibody [19]. This antibody targets an epitope centered on p-tau at serine 202 and threonine 205 [20–22]. However, while phosphorylations at both residues can be measured in biofluids, only p-tau205 has been shown to display a good performance as an AD biomarker [13, 16, 23–25]. Beyond its relationship with neuropathology, p-tau205 distinguishes itself from other soluble p-tau measurements (such as p-tau181, p-tau217, and p-tau231) by exhibiting abnormal concentrations later in the AD *continuum*, closer to symptom onset and tau-PET positivity [13, 14, 26, 27]. As a result, fluid p-tau205 measurements have demonstrated a stronger association with neurofibrillary tau pathology than Aβ plaque pathology (both indexed by PET), a highly distinctive characteristic not observed for p-tau181, p-tau217 or p-tau231 [13, 16, 17]. Because of these characteristic, p-tau205 is central to the ongoing efforts of establishing an *in vivo* fluid biological staging scheme of AD [14, 27–29]. Supportive of this perspective, the Alzheimer’s Association (AA) new criteria for diagnosing and staging AD include this biomarker in the fluid staging framework, with one fluid stage defined by positivity on p-tau205 [29].

Until recently, p-tau205 measurements were exclusively available using mass spectrometry methods [16–18, 23]. However, a recent development introduced the first high-throughput immunoassay capable of measuring p-tau205 in cerebrospinal fluid (CSF) [13, 14], a breakthrough that aims to make this biomarker more widely available in clinical settings and clinical trials. In addition, there is no previous study reporting the longitudinal changes of p-tau205 in sporadic AD. In the current study we firstlyaimed to characterize immunoassay-based CSF p-tau205 measurements across clinical groups and biological categories and to investigate its cross sectional and longitudinal association with pathological AD hallmarks in living patients (using Aβ-PET, tau-PET and MRI) as well as cognitive function. Secondly, we evaluated if CSF p-tau205 measurements can be used for the staging of AD when combined with established biomarkers (CSF Aβ42/40 and p-tau217) and whether the resulting CSF stages can predict longitudinal changes in AD hallmarks, as well as clinical progression to AD dementia.

## METHODS

### Participants

The present study included participants from two independent cohorts, the Swedish BioFINDER-1 (NCT01208675) and BioFINDER-2 (NCT03174938) (Lund University, Lund, Sweden). BioFINDER-2 participants had a more extensive phenotyping with broader number of biomarkers, whereas BioFINDER-1 had a longer follow-up, which allowed accurate tracking of disease progression. Participants recruitment was performed at the Skåne University Hospital and the Hospital of Ängelholm (Sweden). All participants underwent a lumbar puncture with CSF collection at baseline. Participants were stratified as Aβ-negative or -positive (by CSF or PET) cognitively unimpaired (CU- and CU+, respectively), Aβ-positive mild cognitive impairment (MCI+), Aβ-positive AD dementia (AD), cognitively impaired non-AD patients, as Aβ-positive (non-AD+) or -negative (non-AD-; all non-AD-cases in BioFINDER-1 are clinically diagnosed as MCI). Participants’ diagnosis was determined by consensus of memory clinic physicians and a neuropsychologist. MCI diagnosis was determined as a performance of less than 1.5 standard deviation from age and education stratified norms on at least one domain from an extensive neuropsychological battery examining verbal, memory, visuospatial, and attention/executive domains as described previously for BioFINDER-1 [30] and BioFINDER-2 [31]. To be noted, participants with dementia and Aβ−PET scans were not available in the BioFINDER-2 study. AD dementia diagnosis in BioFINDER-2 was determined based on the criteria from the Diagnostic and Statistical Manual of Mental Disorders Fifth Edition and confirmation of Aβ-positivity using CSF biomarkers. This is in line with fulfilling the criteria for probable AD according the International Working Group [32] or AD at stages 4-6 according to the Alzheimer’s Association (AA) criteria [29]. Individuals diagnosed with non-AD cognitive impairment fulfilled the criteria for dementia or minor neurocognitive disorder due to frontotemporal dementia, Parkinson’s disease, dementia with Lewy bodies, vascular dementia, progressive supranuclear palsy, corticobasal syndrome, multiple system atrophy, or primary progressive aphasia as previously described [33]. All participants included in the present study included at least one CSF p-tau205 measurement. All participants provided written informed consent, and ethical approval was granted by the Regional Ethical Committee in Lund (Sweden).

### CSF biomarker measurements

For all BioFINDER-1 and BioFINDER-2 participants, CSF amyloidosis was determined using CSF Aβ42/40 and CSF Aβ42 was measured using the Elecsys β-Amyloid (1–42), electrochemiluminescence immunoassays on a fully automated cobas e 601 instrument (Roche Diagnostics International Ltd., Rotkreuz, Switzerland), whereas CSF Aβ40 concentrations were measured with a robust prototype assay of the Roche NeuroToolKit on cobas e 601 instruments (Roche Diagnostics International Ltd, Rotkreuz, Switzerland). CSF and plasma p-tau-217 levels were measured using an immunoassay developed by Lilly Research laboratories on the Meso-Scale Discovery Platform as previously described [33, 34].

CSF p-tau205 concentrations in BioFINDER-1 and BioFINDER-2 cohorts were determined using an in-house immunoassay developed at the University of Gothenburg (Sweden); all measurements were performed using a Simoa HD-X platform (Quanterix) at Clinical Neurochemistry Laboratory, Sahlgrenska University Hospital, Mölndal (Sweden). CSF p-tau205 Simoa immunoassay development and validation has been previously described [13]. In brief, the CSF p-tau205 immunoassay is composed of a capture antibody selective against phosphorylated tau at serine 205 and an N-terminal tau antibody for detection. Eight-point calibration curves were generated using commercially available GSK-3β phosphorylated recombinant full-length Tau411 (SignalChem) and run in duplicates.. All plates included internal quality control samples, and these were run in duplicates before and after the samples. Repeatability and intermediate precision values in the cohort was ˂15%.

### Imaging measures

Detailed information regarding imaging acquisition and processing for BioFINDER-1 and BioFINDER-2 can be found elsewhere [33, 35]. In BioFINDER-1, Aβ-PET was acquired 90-110 min after the injection of ∼185 MBq [^18^F]flutemetamol. Tau-PET was not available in BioFINDER-1. In BioFINDER-2, Aβ-PET was also acquired using[^18^F]flutemetamol, whereas tau-PET was acquired after 70-90 min post injection of ∼370 MBq [^18^F]RO948. To be noted, most AD dementia participants did not undergo imaging for Aβ-PET as per study design. Neurodegeneration was assessed in both cohorts using cortical thickness from structural magnetic resonance image (MRI) acquired with high resolution T1-weighted anatomical magnetization-prepared rapid gradient echo (MPRAGE) images (1mm isotropic voxels). Volumetric segmentation and parcellation of T1-images were performed using FreeSurfer (v.6.0, https://surfer.nmr.mgh.harvard.edu). For the main analysis, the variables of interest were measured in regions known to be specifically affected in AD. For Aβ-PET, mean Standardized

Uptake Value Ratio (SUVR) was calculated in a neocortical meta-region of interest (ROI) similar to the Centiloid mask using the whole cerebellum as reference region. For tau-PET, mean SUVR was calculated in a temporal meta-ROI for main analyses and an earlier region (medial temporal lobe) [36], in both using the inferior cerebellum as the reference region. Finally, AD-specific cortical thickness meta-ROI included temporal regions with known vulnerability to neurodegeneration in AD, as previously described [37].

### Cognitive measures

In both BioFINDER-1 and BioFINDER-2, cognition and cognitive decline were assessed using two different methods: (i) Mini-mental state examination (MMSE), used as a measure of global cognition and (ii) a modified version of the preclinical Alzheimer’s cognitive composite-5 (mPACC), a more sensitive measure of cognitive decline (especially in early stages) and commonly used in research settings. The latter test was calculated as the average of four z-scores. For tests of memory, the 10-word delayed recall task from the Alzheimer’s Disease Assessment Scale-Cognitive subscale [ADAS-cog] was used, weighted twice, to preserve the weight of memory tests in the original PACC (53), for verbal ability animal fluency was used, for executive function Trail Making Test A [TMT-A], and for global cognition, the MMSE was used.

### Cohort stratification

CSF p-tau205 concentrations in BioFINDER-2 were assessed beyond clinical groups. First, participants were classified according to the presence/absence of Aβ (A, using CSF Aβ42/40 or PET) and tau pathology (T, tau-PET) into AT groups: A-T-, A+T-, A+T+ and A-T+. CSF Aβ42/40 validated cut-offs were specific for each platform (Innotest: 0.752, Elecsys: 0.08, Lumipulse: 0.72, and MSD: 0.752). When CSF was not available in BioFINDER-2, we used Aβ-PET to assess Aβ-status, using previously validated cutoffs (SUVR>1.03 in a neocortical region using whole cerebellum as reference region) [34]. Aggregated tau pathology was dichotomized into positive and negative based on the tau-PET SUVR in the meta-temporal ROI.

Additionally, we stratified participants using the recently proposed AA diagnostic criteria [29]. In BioFINDER-2 Aβ pathology status was determined using Aβ-PET when available (CSF Aβ42/40 ratio was used for dementia cases due to the lack of Aβ-PET measurements in these participants). The medial temporal lobe (MTL) classification using 1.34 as threshold and for the neocortical region, using 1.36 as a threshold. Finally, for the high tau-PET stage, we used the multiblock barycentric discriminant analysis (MUBADA) region, using a previously validated cutoff (SUVR>1.46) [38]. For Braak staging, we used the Braak I-II (SUVR >1.38), Braak III-IV (SUVR>1.36) and Braak V-VI region (SUVR> 1.22).

### Statistical analysis

Differences in CSF p-tau205 levels by groups were assessed using ANCOVA adjusting for age and sex, followed by Tukey’s corrected post hoc pairwise comparisons. We assessed cross-sectional associations between CSF p-tau205 levels and AD biomarkers using linear regression models with AD biomarkers as outcomes and CSF p-tau205 as predictor adjusting for age and sex (and years of education for cognitive outcomes). Separate models were run by Aβ-status groups. Linear mixed models were used to assess the associations between baseline CSF p-tau205 levels and longitudinal AD biomarkers, using random intercepts and time-slopes, adjusting for age and sex (and years of education for cognitive outcomes). Linear mixed models were also used to assess the longitudinal changes of CSF p-tau205 levels by Aβ-status, using random intercepts and time-slopes, adjusting for age and sex. For comparing longitudinal change in CSF p-tau205 with change in other biomarkers, rates of change of CSF p-tau205 levels were calculated using linear mixed models with time as the sole predictor. Similarly, we also calculated rate of change of AD biomarkers. Then, linear regression models were used to assess the association between CSF p-tau205 rates of change and AD biomarkers’ rates of change, adjusting for age and sex (and years of education for cognitive outcomes). LOESS regressions were used to fit the progression of biomarkers abnormalities across continuous measures of pathology (*i.e.,* Aβ- and tau-PET) or CSF stages. For this, all biomarkers were z-scored using information from a control group of CU-individuals.

CSF Aβ42/40, p-tau217 and p-tau205 were dichotomized using a previously validated cutoff for CSF Aβ42/40 and cutoffs extracted from Gaussian Mixture Models for p-tau217 (>11.42) and p-tau205 (>3.13). The interaction of these dichotomized groups was used for creating the CSF staging model in which: CSF stage 1 represents positivity only in CSF Aβ42/40, CSF stage 2 represents also positivity in CSF p-tau217, and CSF stage 3 include participants that were positive in all three biomarkers, while CSF stage 0 includes biomarker-negative individuals. Individuals that did not follow the hierarchical categorization were excluded from the following analyses (BioFINDER-2: 7.4%; BioFINDER-1: 0%). Similar analyses to those used with the continuous CSF p-tau205 levels were performed using the CSF categorization. Next, we examined progression to dementia using Cox proportional hazard models, adjusting for age, sex, and clinical status at baseline using the CSF staging as predictor. Kaplan-Meier curves were used to visualize clinical progression using survival and survminer packages.

All analyses were performed with R (v.4.3.1). A two-sided p value <0.05 was considered statistically significant. FDR correction was applied to account for multiple comparisons when making comparisons among CSF stages. Non-AD individuals were excluded from all analyses having cortical thickness or cognition as outcome for preventing bias. For cross-sectional analyses, we present BioFINDER-2 results as main results due to availability of larger number of biomarkers. For longitudinal analyses, we present BioFINDER-1 results as main analyses for downstream biomarkers available (atrophy and cognitive decline) due to longer follow-up time. Other results are presented in the supplementary results.

## RESULTS

### 1. Participant characteristics

The demographic information of the participants in the two cohorts analysed in this study is presented in Table 1. The BioFINDER-1 cohort was comprised by 705 individuals: 379 were cognitively unimpaired Aβ-negative (CU-), 160 were cognitively unimpaired Aβ-positive (CU+), 101 had MCI and were Aβ-positive (MCI+), and 65 had MCI and Aβ-negative (non-AD-). The average age was 71.8 (SD 5.25), 389 (55.2%) were women, and 262 (37.2%) were *APOE* ε4 carriers. The BioFINDER-2 cohort included a total of 1364 participants, distributed in groups across the AD *continuum*: 457 CU-, 181 CU+, 175 MCI+ and 184 AD dementia, while 367 exhibited other neurodegenerative diseases (non-AD+ and non-AD-). The average age was 69.1(SD 11.7), 680 (49.9%) were women, and 614 (45.0%) were *APOE* ε4 carriers.

**Table 1:** Cohorts’ demographic description. ^1^ 149 participants missing. ^2^ 37 participants missing. ^3^ 11 participants missing. ^4^ 5 participants missing.

| BioFINDER-2 | CU-<br>(N=457) | CU+<br>(N=181) | MCI+<br>(N=175) | ADdem<br>(N=184) | non-AD+<br>(N=78) | non-AD-<br>(N=289) | Overall<br>(N=1364) |
| --- | --- | --- | --- | --- | --- | --- | --- |
| Age | 62.4 (14.3) | 73.5 (8.32) | 73.0 (7.23) | 73.7 (7.49) | 75.5 (5.70) | 70.1 (9.14) | 69.1 (11.7) |
| Women | 266 (58.2%) | 94 (51.9%) | 83 (47.4%) | 108 (58.7%) | 28 (35.9%) | 101 (34.9%) | 680 (49.9%) |
| APOE ε4 carriers<sup>1</sup> | 167 (36.5%) | 82 (45.3%) | 123 (70.3%) | 128 (69.6%) | 43 (55.1%) | 71 (24.6%) | 614 (45.0%) |
| Education<sup>2</sup> | 13.1 (3.42) | 12.9 (3.75) | 12.9 (4.38) | 12.2 (3.94) | 12.6 (4.46) | 12.2 (3.70) | 12.7 (3.80) |
| CSF p-tau205 | 1.96 (0.749) | 3.73 (1.77) | 4.53 (2.20) | 5.76 (2.19) | 3.23 (1.40) | 2.15 (0.847) | 3.15 (2.01) |
| MMSE | 29.0 (1.19) | 28.7 (1.30) | 26.9 (1.90) | 21.0 (4.21) | 24.3 (4.94) | 26.4 (3.32) | 26.8 (3.79) |

| BioFINDER-1 | CU-<br>(N=379) | CU+<br>(N=160) | MCI+<br>(N=101) | non-AD-<br>(N=65) | Overall<br>(N=705) |
| --- | --- | --- | --- | --- | --- |
| Age | 71.7 (5.13) | 73.0 (5.11) | 72.1 (5.08) | 69.4 (5.70) | 71.8 (5.25) |
| Women | 233 (61.5%) | 88 (55.0%) | 48 (47.5%) | 20 (30.8%) | 389 (55.2%) |
| APOE ε4 carriers<sup>3</sup> | 84 (22.2%) | 91 (56.9%) | 75 (74.3%) | 12 (18.5%) | 262 (37.2%) |
| Years of education<sup>4</sup> | 12.3 (3.33) | 11.9 (3.71) | 11.5 (3.42) | 10.5 (3.08) | 11.9 (3.44) |
| CSF p-tau205 | 1.77 (0.549) | 3.17 (1.54) | 4.32 (1.74) | 1.83 (0.610) | 2.46 (1.44) |
| MMSE | 29.1 (1.04) | 28.8 (1.23) | 26.5 (2.01) | 27.8 (1.90) | 28.5 (1.63) |

### 2. CSF p-tau205 increases across the AD continuum and correlates with imaging biomarkers and cognition

In BioFINDER-2, CSF p-tau205 was significantly increased in all Aβ-positive groups compared with the Aβ-negative groups (CU- and non-AD-; *p*<0.001 for all). This increase was progressive across the AD *continuum* (Figure 1A, Supplementary Table 1). Similar findings were observed in BioFINDER-1 (Supplementary Figure 1A). When classified into AT groups based on Aβ pathology (A, CSF Aβ42/40) and tau pathology (T, tau-PET), CSF p-tau205 increased from A-T-to A+T-(FC=1.70, *p*<0.001), and from A+T-to A+T+ (FC=1.62, *p*<0.001; Figure 1B, Supplementary Table 2). When participants were classified using tau-PET Braak stages or the AA imaging-based staging criteria, CSF p-tau205 increased progressively along with the advance in pathology status (*p*<0.001, for all; Figure 1C-D, Supplementary Tables 3-4).

**Figure 1.**
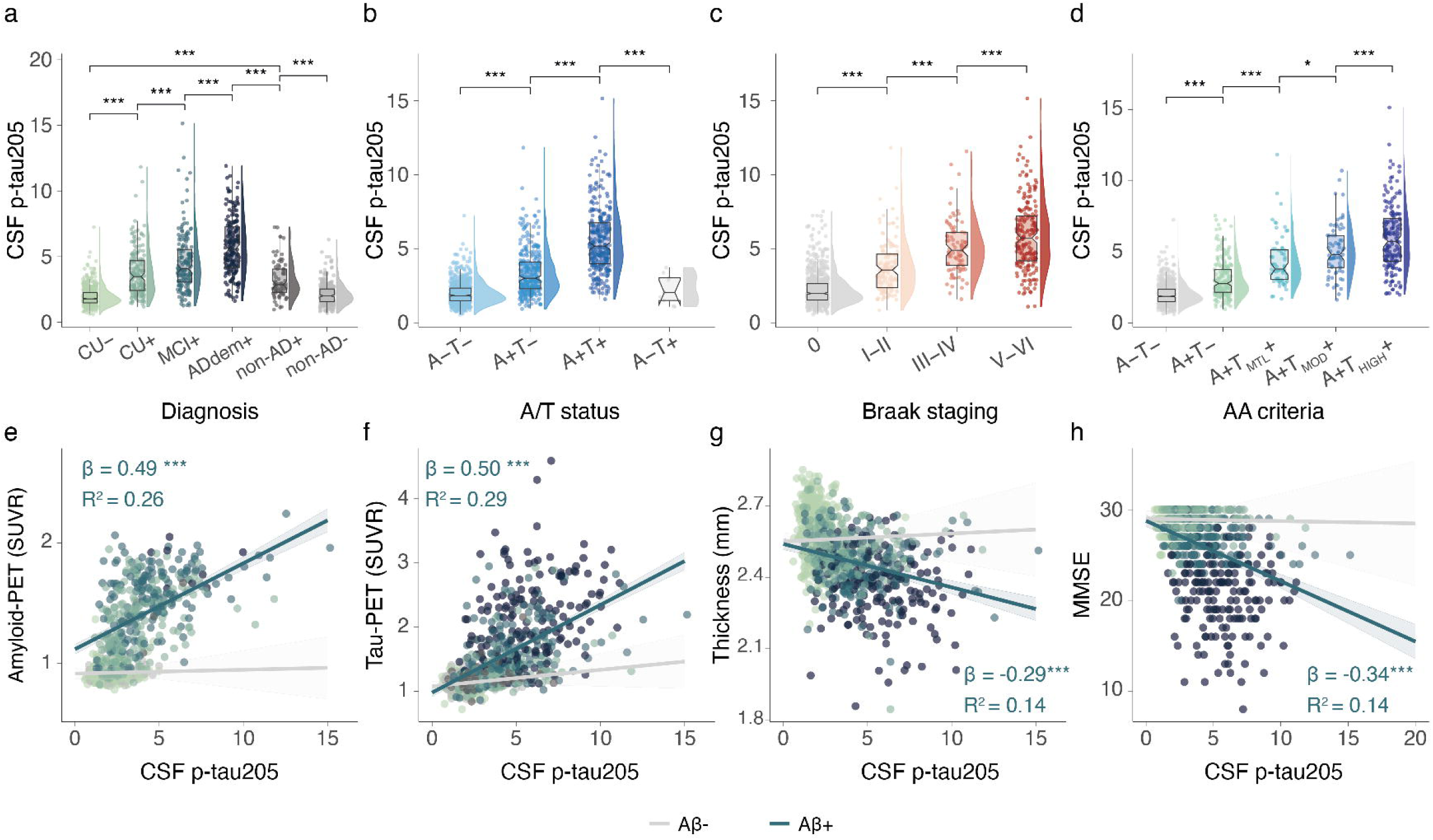
CSF p-tau205 concentrations in BioFINDER-2 by clinical diagnosis (**A**), A/T status (**B**), tau-PET Braak stages (**C**), and AA criteria using tau-PET (**D**). Cross-sectional associations between CSF p-tau205 concentrations and Aβ-PET (**E**), tau-PET (**F**), cortical thickness (**G**) and MMSE (**H**) in BioFINDER-2. Differences in CSF p-tau205 levels by groups were determined using ANCOVA and Tukey’s method for *post-hoc* comparisons. Age and sex (and years of education for cognitive outcomes) were used as covariates in all cases. Aβ (A) status was assessed using CSF Aβ42/40 levels or PET and tau (T) status using tau-PET SUVR based on previously validated cut-offs. Participants with available tau-PET imaging were stratified according to the PET Braak stages in a hierarchical manner, based on regional SUVR cut-offs. *: *p*<0.05; **: *p*<0.01; ***: *p*<0.001. Box plots include all participants, displaying the median and the interquartile range; whiskers show the lower value of maximum/minimum value or 1.5 interquartile range from the hinge. Linear regressions with CSF p-tau205 as predictor and PET, MRI and MMSE as outcome were used to measure the association. Non-AD individuals were excluded from analyses with cortical thickness or cognition. Standardized β (β_std_) and p-values of the associations as well as the R^2^ of the model for the Aβ-positive individuals are shown in the plots. *: *p*<0.05; **: *p*<0.01; ***: *p*<0.001.

Furthermore, CSF p-tau205 significantly correlated with Aβ-PET in Aβ+ individuals in BioFINDER-2 (R^2^=0.26, β[95%CI]=0.49 [0.40, 0.58], *p*<0.001) (Figure1E). The same was true for medial temporal (MTL) (R^2^=0.32, β[95%CI]=0.56 [0.5, 0.63], *p*<0.001) and neocortical (NeoT) tau-PET (R^2^=0.29, β[95%CI]=0.5 [0.43, 0.57], *p*<0.001) (Figure 1F, Supplementary Table 5). Finally, in Aβ+ individuals, CSF p-tau205 was significantly associated with cortical thickness (R^2^=0.14, β[95%CI]= −0.29 [−0.37, −0.21], *p*<0.001) as well as with cognition determined with MMSE (R^2^=0.14, β[95%CI]= −0.34 [−0.42, −0.26], *p*<0.001) (Figure 1G-H) and mPACC5 (R^2^=0.14, β[95%CI]= −0.33 [−0.41, −0.24], *p*<0.001) (Supplementary Table 5). Cross-sectional association with imaging biomarkers and cognitive status was replicated in BioFINDER-1 (tau-PET was not available in BioFINDER-1; Supplementary Figure 1 and Supplementary Table 5).

### 3. Baseline CSF p-tau205 predicts longitudinal imaging biomarkers and cognition

We also investigated whether CSF p-tau205 measurements are useful when predicting AD progression. In BioFINDER-2, higher baseline concentrations of CSF p-tau205 were associated with overtime increased in Aβ and tau-PET tracer uptake (Aβ-PET: R^2^=0.44, β[95%CI]=0.04 [0.03, 0.05], *p*<0.001; MTL Tau-PET: R^2^=0.50, β[95%CI]=0.05 [0.04, 0.06], *p*<0.001; NeoT Tau-PET: R^2^=0.33, β[95%CI]=0.12 [0.1, 0.14], *p*<0.001; Figure 2A-B; Supplementary Table 6). In BioFINDER-1, higher baseline concentrations of CSF p-tau205 were associated with steeper decreases in cortical thickness (R^2^=0.24, β[95%CI]= −0.17 [−0.19, −0.14], *p*<0.001) (Figure 2C) and a more pronounced cognitive decline (mPACC: R^2^=0.38, β[95%CI]= −0.32 [−0.36, −0.28], *p*<0.001; MMSE: R^2^=0.36, β[95%CI]= −0.4 [−0.45, −0.36], *p*<0.001; Figure 2D; Supplementary Table 6).

**Figure 2.**
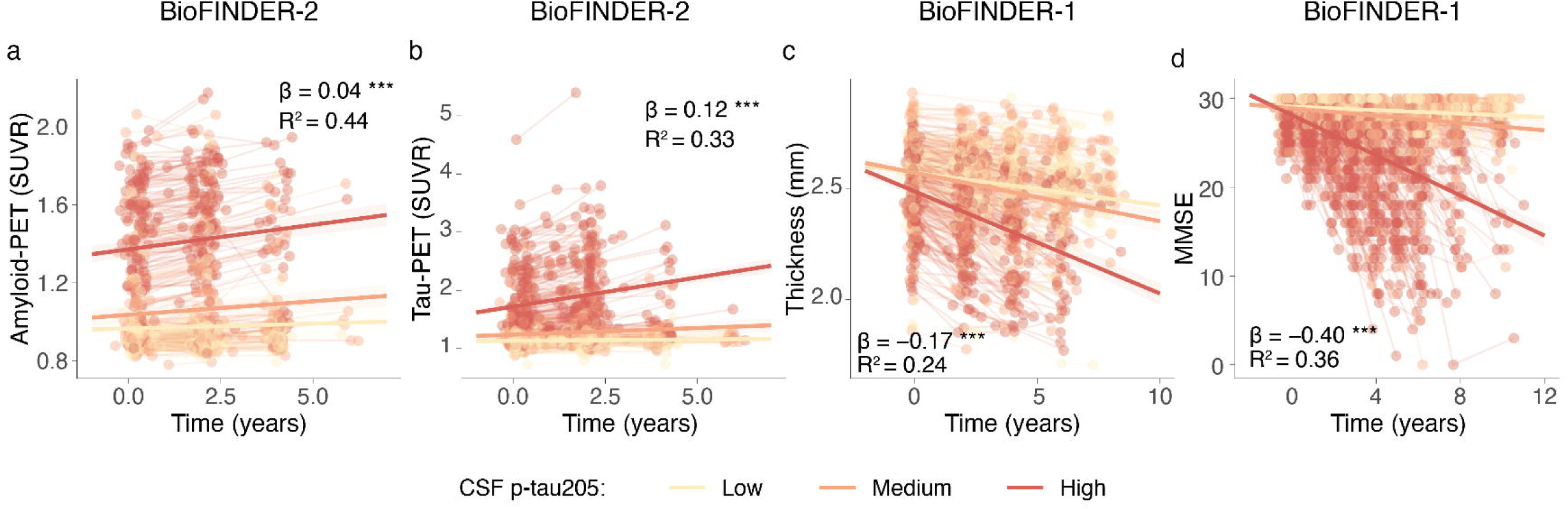
Baseline CSF p-tau205 association with longitudinal Aβ-PET, tau-PET, neurodegeneration, and cognition. Association with longitudinal Aβ-PET (**A**) and tau-PET (**B**) were determined in BioFINDER-2. Association with longitudinal cortical thickness (**C**), and MMSE (**D**) were determined in BioFINDER-1. Non-AD individuals were excluded from analyses with cortical thickness or cognition. We used linear mixed models with PET (SUVR), cortical thickness (mm), and cognitive measures as outcome and the interaction of baseline CSF p-tau205 and time as predictor with random intercepts and random time-slopes. Age and sex (and years of education for cognitive outcomes) were used as covariates. Dots and thin lines represent individual timepoints and trajectories, respectively, for each participant. Each participant is coloured based on its baseline CSF p-tau205 levels. Thick lines and shaded areas represent the mean trajectory over time of each group of CSF p-tau205 baseline levels and its 95%CI. Standardized β (β_std_) and *p*-values of the associations as well as the R^2^ of the model are shown in the plots. Standardized β (β_std_) and *p*-values of the associations as well as the R^2^ of the model are shown in the plots. *: *p*<0.05; **: *p*<0.01; ***: *p*<0.001.

### 4. Longitudinal CSF p-tau205 associate with over time changes in imaging biomarkers and cognition

In BioFINDER-1, we examined the longitudinal changes in CSF p-tau205, and whether they were dependent on Aβ pathology status at baseline in all participants, as well as within CU and CI participants (up to 10 and 8 years of follow up, respectively). Across all subjects, CSF p-tau205 increased longitudinally in Aβ-positive individuals in comparison with those Aβ-negative (time × Aβ-status interaction: β[95%CI]=0.14[0.11, 0.18], *p*<0.001) (Figure 3A). The same was observed in CU (time × Aβ-status interaction: β[95%CI]=0.16[0.12, 0.21], *p*<0.001) and CI participants (time × Aβ-interaction: β[95%CI]=0.10[0.04, 0.16], *p*<0.001; Figure 3B-C and Supplementary Table 7).

**Figure 3.**
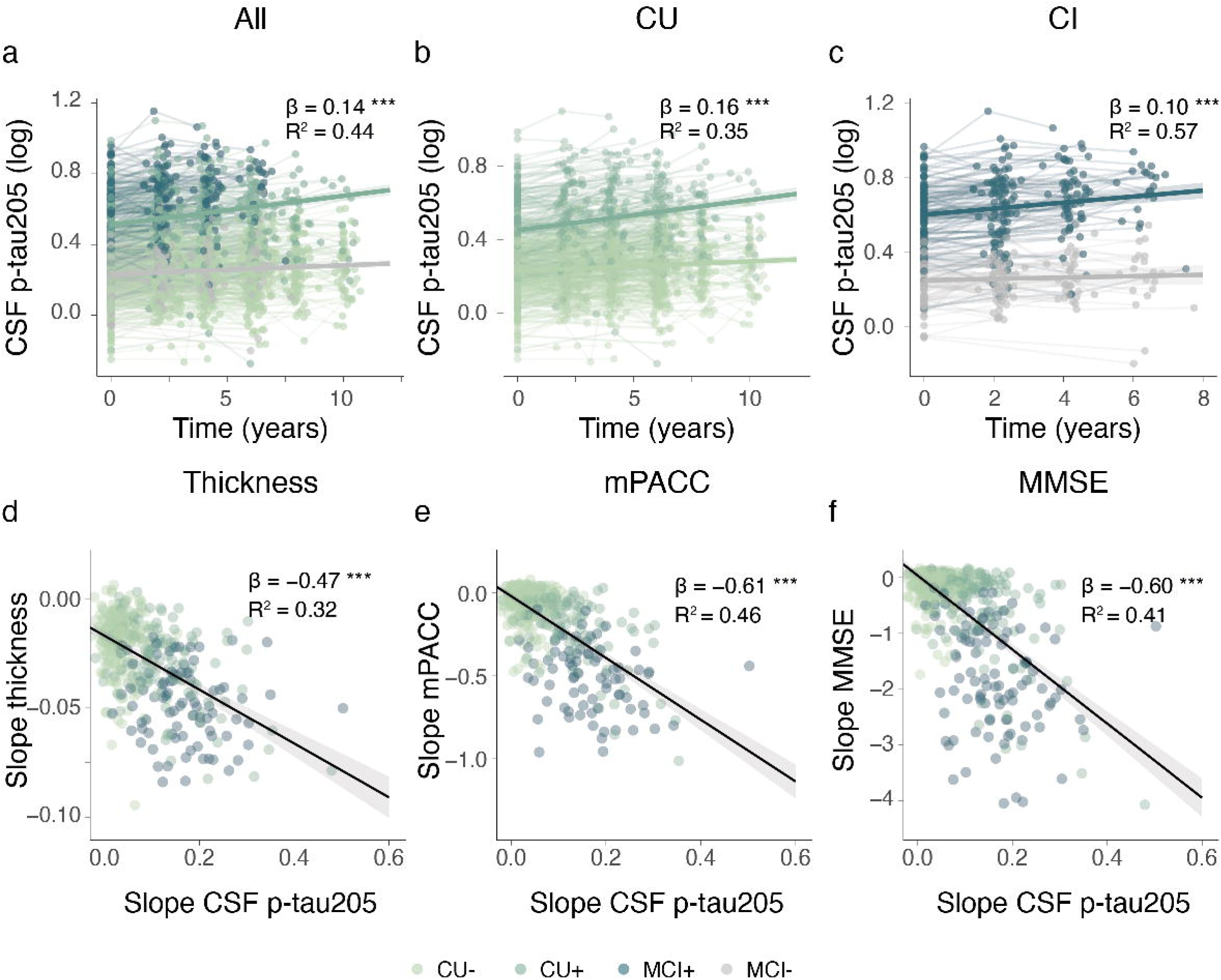
Longitudinal CSF p-tau205 changes classified by baseline Aβ status in all individual (**A**), as well as cognitively unimpaired (**B**) and impaired participants (**C**) from BioFINDER-1. CSF p-tau205 levels were used as outcome in linear mixed models with the interaction between Aβ status and time as predictor. Age and sex were included as covariates. Aβ-status was based on CSF Aβ42/40 levels. Standardized β (β_std_) and p-values of the interaction term are shown in the plots. *: *p*<0.05; **: *p*<0.01; ***: *p*<0.001. Associations between longitudinal CSF p-tau205 changes and longitudinal changes in cortical thickness (**D**) and cognition (**E**, MMSE; **F**, mPACC) in BioFINDER-1. Rates of change of CSF p-tau205 and AD biomarkers levels were calculated using linear mixed models with time as the sole predictor, in independent models. Linear regression models were used to assess the association between CSF p-tau205 rates of change and AD biomarkers’ rates of change, adjusting for age and sex (and years of education for cognitive outcomes).

Subsequently, we investigated the association between longitudinal CSF p-tau205 level changes with over time changes in cortical thickness and cognition. In BioFINDER-1, longitudinal changes in CSF p-tau205 were significantly associated with longitudinal decreases in cortical thickness (R^2^=0.32, β[95%CI]= −0.47 [−0.54, −0.4], *p*<0.001) (Figure 3D). Likewise, longitudinal changes in CSF p-tau205 were associated with overtime worsening in MMSE (R^2^=0.41, β[95%CI]= −0.6 [−0.66, −0.54], *p*<0.001) and mPACC scores (R^2^=0.46, β[95%CI]= −0.61 [−0.68, −0.55], *p*<0.001; Figure 3E-F; Supplementary Table 8).

### 5. CSF p-tau205 in combination with Aβ42/40 and p-tau217 can stage AD

We then investigated how CSF p-tau205 measurements, combined with established CSF fluid biomarkers such as Aβ42/40 and p-tau217, may contribute to AD staging. Cumulative abnormality in fluid biomarkers measurements followed the expected biomarker sequence (that is Aβ42/40, followed by p-tau217, and lastly p-tau205), both when using Aβ-PET and tau-PET uptake as a proxy of AD progression. Notably, the trajectory of CSF p-tau217 levels reached abnormality closer to Aβ-PET positivity threshold when compared to CSF p-tau205 (Figure 4A). Contrarily, CSF p-tau217 became abnormal prior to the tau-PET cutoff for positivity, whereas CSF p-tau205 and neocortical tau-PET become abnormal almost simultaneously (Figure 4B). Thus, the resulting model was defined by four stages: stage-0 (all CSF biomarkers negative), stage-1 (Aβ42/40 positive), stage-2 (Aβ42/40 and p-tau217 positive), and stage-3 (Aβ42/40, p-tau217, and p-tau205 positive) (Figure 4C).

**Figure 4.**
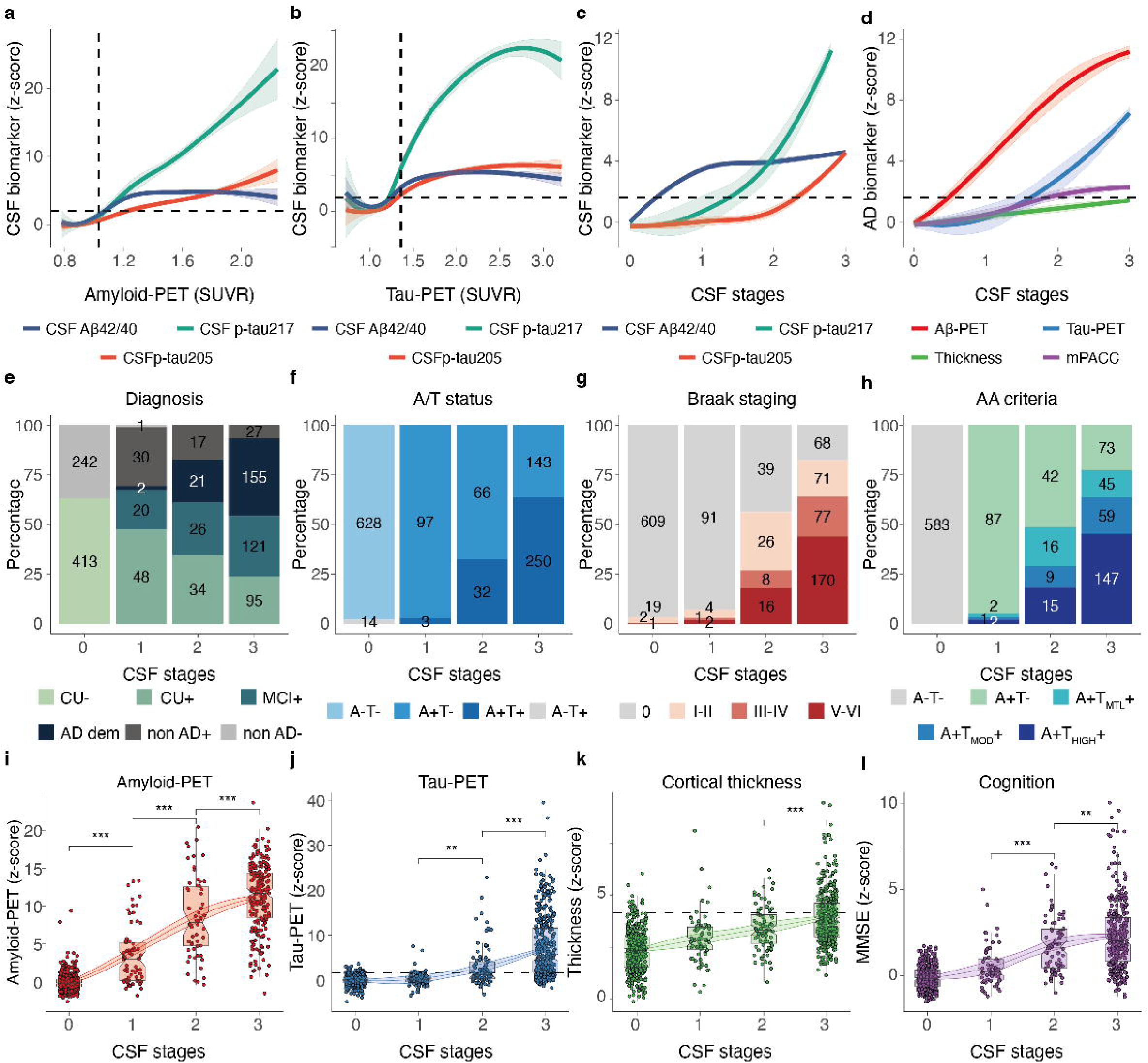
CSF Aβ42/40, p-tau217, and p-tau205 levels and trajectories across Aβ-PET and tau-PET SUVRSs in BioFINDER-2 (**a-b**). The scatterplots depict CSF biomarker values (z-scores) in relation to Aβ-PET (**a**) and tau-PET (**b**) SUVRs, used as a proxy of AD pathology progression. The line represents the locally estimated scatterplot smoothing (LOESS) regression. Biomarker thresholds for positivity are indicated by the vertical dashed lines. Horizontal dashed lines represent z-score=1.96 (95%CI of the control group). CSF Aβ42/40, p-tau217, and p-tau205 levels (**c**); and Aβ-PET, tau-PET, cortical thickness and MMSE (**D**) across CSF stages. CSF stages: stage-1 (positive Aβ42/40), stage-2 positive (Aβ42/40 and p-tau217), and stage-3 (positive Aβ42/40, p-tau217, and p-tau205). CSF staging across diagnostic groups (**e**), A/T status (**f**) tau-PET defined Braak stages (**g**), AA criteria using tau-PET (**h**) in BioFINDER-2. *For AA criteria*: Aβ pathology status was determined using Aβ-PET when available (CSF Aβ42/40 ratio was used for dementia cases due to the lack of Aβ-PET measurements in these individuals). Tau-PET was used to determined tau pathology status and classified into: T-(no tau-PET uptake), T_MTL+_ (tau-PET uptake restricted to medial temporal areas), T_MOD+_ (tau-PET uptake in the moderate SUVR range on neocortex), and T_HIGH+_ (neocortical tau-PET uptake in the high SUVR range). Stacked bar charts represent the percentages of CSF stage-0, stage-1, stage-2 and stage-3 participants (exact number of participants is displayed within the bar plot). Cross-sectional levels of amyloid-(**i**) and tau-PET burden (**j**), cortical thickness (**k**) and MMSE (**l**) across CSF stages in BioFINDER-2 participants. AD biomarker values were z-scored based on a group of CU-participants, and all increases represent increase in abnormality. Coloured lines and bands represent the LOESS regression and its 95% CI. *: *p*<0.05; **: *p*<0.01; ***: *p*<0.001.

We subsequently investigated the distribution of diagnostic and pathological classifications across the defined CSF stages (Figure 4, Supplementary Table 9). In BioFINDER-2 participants, higher CSF stages were associated with more advanced clinical and pathological stages (Figure 4E-H). Notably, positivity for CSF p-tau205 (that is CSF stage-3) mainly included individuals with (i) MCI+ and AD diagnoses, (ii) intermediate to advanced tau-PET Braak stages (Braak III-IV and V-VI), (iii) positive for Aβ-PET and tau-PET (A+T+), and (iv) advanced AD according to the AA staging criteria using imaging biomarkers (A+T_MTL_+, A+T_MOD_+ and A+T_HIGH_+). In this last case, tau-PET abnormality across CSF stages revealed that while medial temporal tracer uptake was clearly abnormal for most cases at CSF stage-2, advanced tracer binding in neocortical was a more characteristic feature of participants classified as positive for CSF p-tau205, that is CSF stage-3.

We then explored the association between the defined CSF stages with Aβ pathology (Aβ-PET), tau pathology (tau-PET), neurodegeneration (cortical thickness in AD signature areas), and cognitive status (MMSE) (Figures 4I-K. Although each biomarker followed a distinct trajectory, the magnitude of abnormalities across all assessed AD hallmarks exhibited a consistent increase with advancing CSF stages. Abnormal mean levels of Aβ-PET tracer binding were already evident at CSF stage-1, whereas for tau-PET these become abnormal at CSF stage-2. On the other hand, abnormal measurements of cortical thickness appeared to be more defining aspects of CSF stage-3, with approximately 50% of the individuals showing z-score above 2SD. Finally, mean z-scores in MMSE crossed the threshold already at CSF stage-2, but these were substantially more overt at CSF stage-3. Changes in imaging biomarkers as well as cognition across the fluid stages are summarised in Figure 4D.

### 6. Immunoassay-based CSF stages predict disease and clinical progression

We assessed the utility of CSF p-tau205 measurements within the framework of CSF staging to predict longitudinal outcomes, including brain atrophy (cortical thickness), and cognitive decline (measured by MMSE and mPACC) in BioFINDER-1 (Figure 5, Supplementary Table 10). Baseline CSF stage-2 and 3 were associated with significant decreases in cortical thickness (β[95%CI] stage 2= −0.11, *p*=0.014; β[95%CI] stage 3= −0.38, *p*<0.001). Similar findings, but with more pronounced slopes, were observed for baseline CSF stage-2 and CSF stage-3 when investigating cognitive decline measured by mPACC (β[95%CI] stage 2= −0.25, *p*<0.001; β[95%CI] stage 3= - 0.83, *p*<0.001) and MMSE (β[95%CI] stage 2= −0.39, *p*<0.001; β[95%CI] stage 3= −0.95, *p*<0.001). These results were replicated and consistent in the BioFINDER-2 cohort (Supplementary Table 10).

**Figure 5.**
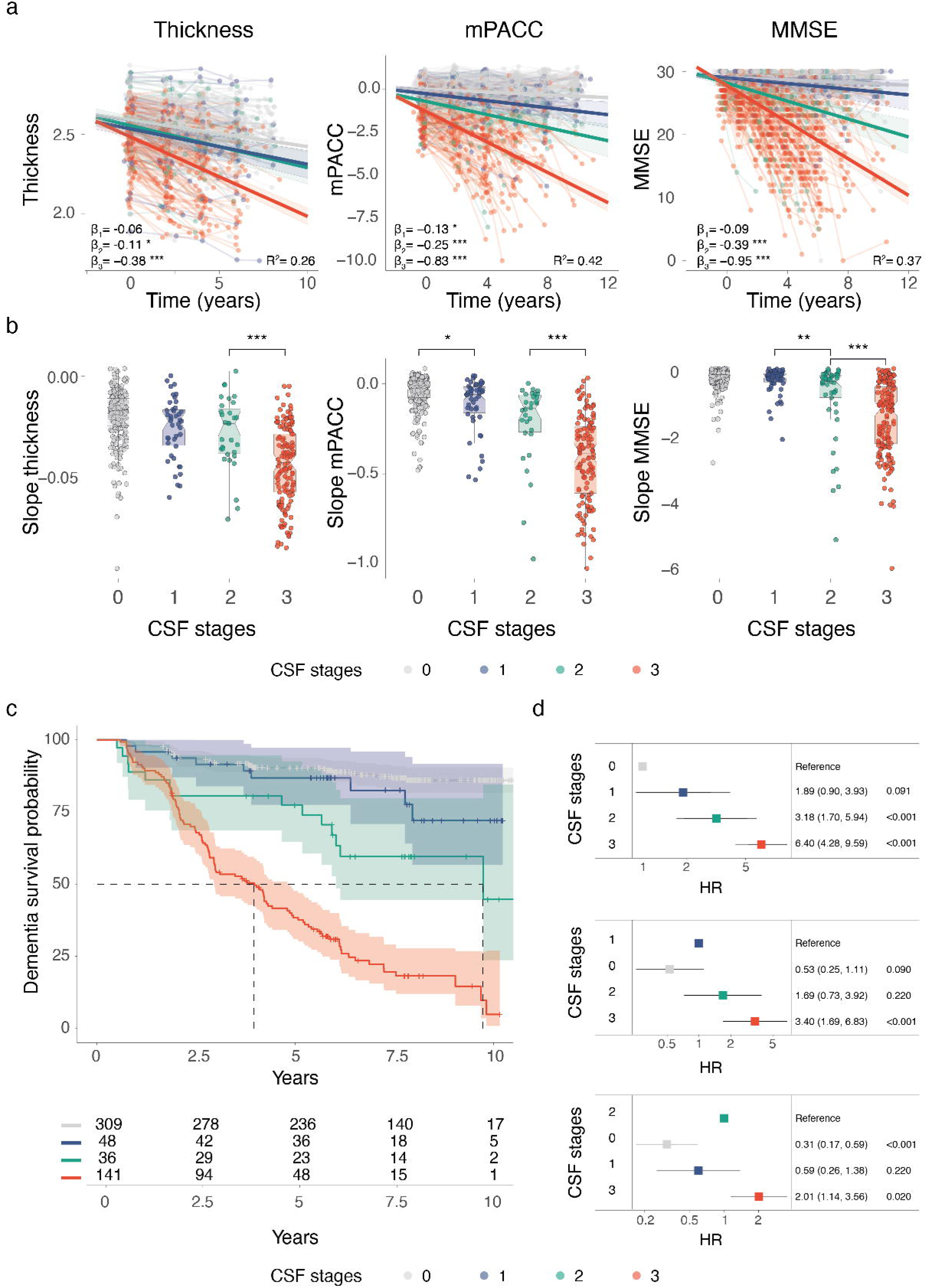
Baseline CSF stages association with longitudinal changes in cortical atrophy, MMSE, and mPACC, determined in BioFINDER-1 (**A-B**). We used linear mixed models with cortical thickness (mm) and cognitive measures as outcome and the interaction of baseline CSF stages and time as predictor with random intercepts and random time-slopes. Age and sex (and education for cognitive outcomes) were used as covariates. Dots and thin lines represent individual timepoints and trajectories, respectively, for each participant. Each participant is coloured based on its baseline CSF staging group. Thick lines and shaded areas represent the mean trajectory over time of each CSF staging group at baseline and its 95%CI (A). Standardized β (β_std_) and *p*-values of the associations as well as the R^2^ of the model are shown in the plots. Individual slopes for participants are shown in **B** in boxplots by CSF stage. Progression to dementia by baseline CSF stage is shown in C-D. Survival curves for progression to dementia in the CSF stages, including a table of total number of participants available at each time point. The dashed line in a indicates the time point at which 50% of a group had progressed to dementia. (**D**) Forest plots showing the HRs and 95% CIs derived from the survival analyses shown in (**C**), from Cox regression models including age, sex, and cognitive stage at baseline as covariates using different CSF stages as reference group. *: *p*<0.05; **: *p*<0.01; ***: *p*<0.001. CSF stages: stage-1 (positive Aβ42/40), stage-2 positive (Aβ42/40 and p-tau217), and stage-3 (positive Aβ42/40, p-tau217, and p-tau205).

In addition, in BioFINDER-2 we also evaluated the association between CSF stages and longitudinal Aβ and Tau-PET (Supplementary Table 10). Aβ-positive CSF stages (that is stage-1, −2 and −3) were all associated with longitudinal increases in Aβ-PET tracer binding (*p*<0.001). The same was true for longitudinal tau-PET uptake in early brain regions (MTL) (*p*<0.001). However, only baseline CSF stage-2 and −3 associated with overtime changes in global tau-PET tracer binding (NeoT), with CSF stage-3 displaying the most pronounced longitudinal increases.

Lastly, we examined the utility of p-tau205 measurements within the CSF staging model for predicting clinical progression (Figures 5C and 5D, Supplementary Table 11). Specifically, we evaluated the model’s ability to predict progression from baseline CU or MCI to AD dementia in the BioFINDER-1 cohort, with up to 10 years of follow-up after the baseline visit. The risk of progression to AD dementia (progressors, n=171) was highest among individuals classified as CSF stage 3 (HR=6.40 [4.28, 9.59], *p*<0.001), followed by those at stage 2 (HR=3.18 [1.70, 5.94], *p*<0.001; Figure 6D, first row). Notably, individuals at CSF stage 3 also had a significantly higher risk of progression compared to those at stage 2 (HR=2.01 [1.14, 3.56], *p*<0.05; Figure 6D, last row).

## DISCUSSION

In the present study, we characterized CSF p-tau205 biomarker cross-sectionally and longitudinally, and evaluated its performance, both as a standalone biomarker and when combined with established CSF biomarkers into a fluid staging system. Our findings support p-tau205 as an important biomarker milestone in the natural history of AD, which more accurately reflects early aggregated tau pathology. Our findings demonstrate that CSF p-tau205 biomarker measurements are tightly linked with tau-PET changes in AD, increase progressively across AT and tau-PET defined classifications, and are significantly associated with *in vivo* AD pathological hallmarks and cognition. We also show that high baseline levels of CSF p-tau205 are associated with over time increases in plaque and tangle deposition as well as accelerated cortical atrophy and worsened cognition. Moreover, longitudinal changes in CSF p-tau205 were associated with progressive neurodegeneration and cognitive decline. Lastly, we demonstrate that CSF p-tau205, when combined with other immunoassay-based biomarkers (CSF Aβ42/40 and p-tau217), can be used for staging AD. In this staging system, individuals classified as higher CSF stages at baseline displayed a more severe worsening in cortical thickness and cognitive decline and exhibited a higher likelihood of progressing to AD dementia compared with those at lower CSF stages. Importantly, this validation was performed using a scalable immunoassay, facilitating the translation of previous mass spectrometry-based findings [17, 26, 27] into a tool suitable for clinical practice to support biological staging of AD and prediction of cognitive decline.

Phosphorylation at threonine 205 represents an immunohistochemical signature of tau aggregation and deposition into neurofibrillary tangles [39]. Therefore, quantifying this critical milestone in AD pathophysiology, particularly through immunoassay platforms, may significantly improve ongoing efforts to develop accessible and cost-effective analytical methods for detecting underlying tau pathology in living AD patients. The cross-sectional findings presented here aligned well with preceding CSF p-tau205 literature using mass spectrometry [15–17, 24, 26]. First, increased levels of CSF p-tau205 concentrations are highly specific for AD. As previously shown [13, 14], CSF p-tau205 levels were already abnormal in asymptomatic AD patients, gradually increased as a function of clinical severity. Moreover, while this biomarker was elevated in CSF Aβ42/40 positive individuals with normal tau-PET tracer uptake (A+T-), CSF p-tau205 levels seemed tightly linked with the underlying status of tau pathology, as demonstrated by the progressive biomarker concentration across tau-PET defined classifications (Braak stages and MTL/NeoT tau-PET status).

Previous cross-sectional data showed that CSF p-tau205 levels correlated with Aβ and tau pathology indexed by PET, as well as neurodegeneration determined by MRI and cognitive performance [13]. The present study not only corroborates these preceding findings in independent and larger cohorts but also extends them by providing the first longitudinal assessment of p-tau205 in sporadic AD. Specifically, elevated baseline CSF p-tau205 levels predicted longitudinal increases in tau-PET burden, as well as worsening neurodegeneration and cognitive decline. Furthermore, longitudinal changes in CSF p-tau205 were higher in those individuals who were Aβ-positive and were associated with increasing tau-PET uptake, decreasing cortical thickness, and cognitive decline.

In recent years, soluble p-tau205 has gained significant attention in the AD biomarker field [13, 14, 16, 17, 24, 26–29]. This specific phosphorylation plays a key role in misfolded tau deposition and is central to the neuropathological definitive diagnosis of AD. Moreover, as a CSF biomarker, p-tau205 emerges at a distinct time point across the AD *continuum* compared to other soluble p-tau forms [14, 26, 27, 29] and exhibits stronger correlation with tau-PET than other p-tau species [16, 17]. These characteristics have resulted in p-tau205 being suggested as a potential candidate for the biological staging of AD using fluid biomarker [28, 29]. The clinical utility of p-tau205 as a staging biomarker of AD has been recently explored using MS measurements of p-tau205 [27]. The present study also investigated whether this may also be achievable through immunoassay-based staging. For this purpose, we firstly explored if levels of CSF p-tau205 do indeed increase after CSF p-tau217, as mass spectrometric reports have previously demonstrated [14, 26, 27]. Thus, we compared the trajectory of CSF p-tau217 as p-tau205 in CSF Aβ42/40 positive cases using Aβ-PET and tau-PET as a surrogate indicators of AD progression. Our findings show that CSF Aβ42/40 is the first emerging biomarker, followed by CSF p-tau217 and later by CSF p-tau205, both across Aβ-PET and tau-PET, altogether suggesting the suitability of this novel assay for AD staging. Notably, we observed that the trajectory of CSF p-tau205 intersected neocortical tau-PET positivity threshold when fluid biomarker levels were abnormally elevated (unlike CSF p-tau217, which become abnormal before both CSF p-tau205 and neocortical tau-PET). In other words, CSF p-tau205 and neocortical tau-PET seemingly become positive simultaneously Fluid stages successfully stratified increasing severity in Aβ pathology, tau pathology and neurodegeneration. Abnormal mean Aβ-PET levels were already observed at stage-1, a fluid stage characterized by positivity in CSF amyloid (but not CSF p-tau). However, the trajectory and mean levels of tau-PET across CSF stages was different across different brain regions. First, while abnormal mean tracer uptake in medial temporal regions was already present at CSF stage-2, this was a more prominent feature of CSF stage-3 or CSF p-tau205 positive individuals. This was more evident when examining neocortical and Braak V-VI, with abnormal mean tracer binding representing a clear feature of CSF stage-3Specifically, CSF stage-3 comprised more than 50% of participants at intermediate-to-advanced clinical and pathological categories, namely MCI+ or AD diagnoses, A+T+, tau-PET defined Braak Stages III-IV or V-VI, and A/MTL/Neo+ or A/MTL/Neo++. Notably, CSF stage-3 also included a comparatively smaller percentage of participants typically associated with early classifications of disease progression and severity, such as CU+, A+T-, A+ or A/MTL+, and Braak stages I-II. Thus, this suggests that increased CSF p-tau205 represents an important biomarker landmark in AD progression [28, 29] and contributes to the idea that high levels of this marker could be useful to further characterize individuals across the AD *continuum*.

An important contribution of the present study is assessing whether using CSF p-tau205 for staging individuals at baseline can predict longitudinal changes in Aβ pathology, tau pathology, cortical thickness and cognition. Except for Aβ-PET, being classified at baseline as higher fluid stage was associated with longitudinal increases in tau-PET tracer uptake over 6 years, as well as steeper decreases in both cortical thickness over 6 to 10 years. The later was also true for cognition (over 6 to 12 years), both when using a more global or more sensitive test (MMSE or mPACC, respectively). These findings suggest that the utility of determining CSF p-tau205 abnormality expands beyond the biomarker phenotyping and disease staging of AD individuals, by predicting the overtime tangle burden accumulation, neurodegeneration and cognitive decline. The present immunoassay-based staging framework was also effective in predicting progression to dementia. Nearly 100% of individuals in stage 3 developed AD dementia within 10 years, whereas for stage 2 (Aβ and p-tau217 positive), it was only 50%. This suggests that individuals who are p-tau205 positive are at a significantly higher risk of progressing to AD dementia in the following years.

To our knowledge, this is the first study to evaluate the longitudinal changes in p-tau205 levels in sporadic AD. These findings have important implications for clinical trial screening and patient management. Notably, anti-amyloid therapies have shown the greatest efficacy in amyloid-positive individuals with low tau-PET burden [40]. Our results indicate that changes in CSF p-tau205 abnormality occur simultaneously with changes in neocortical tau-PET abnormality, suggesting that p-tau205 levels could serve as a useful proxy for tau-PET burden and aid in therapeutic decision-making. Furthermore, the ability of p-tau205 to predict disease progression is valuable for ensuring that patients receiving specific treatments are at a high risk of developing AD, thereby optimizing therapeutic strategies. Future studies should explore whether longitudinal changes in p-tau205 levels could also be used to monitor treatment effects.

This study has several strengths, including the use of an immunoassay platform for measuring CSF p-tau205, the inclusion of two independent cohorts, and the incorporation of longitudinal data to assess biomarker dynamics over time. However, certain limitations should be acknowledged. In the BioFINDER-1 cohort, the absence of individuals with dementia and the lack of tau-PET data limits the ability to evaluate biomarker performance across the full AD spectrum in this cohort. Additionally, in the BioFINDER-2 cohort, the availability of longitudinal data was limited, which may restrict the validation of p-tau205’s temporal progression in this cohort.

## CONCLUSIONS

To conclude, in the current work we validated CSF p-tau205 as a biomarker that emerges later in the AD *continuum* when compared with other p-tau forms, and could be used as an accessible alternative to tau PET to track and predict underlaying tau pathology in AD. Our findings highlight the potential of immunoassay-based measures for staging AD, not only in identifying AD but, more importantly, in predicting longitudinal changes in AD neuropathological hallmarks and cognition. Furthermore, by incorporating CSF p-tau205 into an AD staging framework, we demonstrate that positivity for this biomarker can be a valuable indicator of disease progression.

## ABBREVIATIONS

AA: Alzheimer’s Association
Aβ: Amyloid-β
A-T-: Aβ and tau negative
A+T-: Aβ-positive tau negative
A+T+: Aβ and tau positive
A-T+: Aβ-negative tau positive
AD: Alzheimer’s disease
βstd: Standardized β
CI: Cognitively impaired
CI-: Cognitively impaired Aβ-negative
CI+: Cognitively impaired Aβ-positive
CI95%: Confidence interval
CSF: Cerebrospinal fluid
CU: Cognitively unimpaired
CU-: Cognitively unimpaired Aβ-negative
CU+: Cognitively unimpaired Aβ-positive
HR: Hazard ratio
MCI: Mild cognitive impairment
MCI-: Mild cognitive impairment Aβ-negative
MCI+: Mild cognitive impairment Aβ-positive
MMSE: Mini-mental state examination
mPACC: Modified preclinical Alzheimer’s cognitive composite
MRI: Magnetic resonance imaging
MTL: Medial temporal lobe
MTBR: Microtubule-binding region
MUBADA: multiblock barycentric discriminant analysis
NeoT: Neocortical
NFTs: Neurofibrillary tangles
NIA-AA: National Institute of Aging and Alzheimer’s Association
Non-AD-: Non-Alzheimer’s type dementia Aβ-negative
Non-AD+: Non-Alzheimer’s type dementia Aβ-positive
PET: Positron emission tomography
P-tau: Phosphorylated tau
ROI: Region of interest
SUVR: Standardized uptake value
TMT-A: Trail Making Test A

## DECLARATIONS

### Ethics approval and consent to participate

All participants gave written informed consent, and ethical approval was granted by the Regional Ethical Committee in Lund, Sweden.

### Consent for publication

Not applicable.

### Availability of data and materials

Anonymized data will be shared by request from a qualified academic investigator for the sole purpose of replicating procedures and results presented in the article and as long as data transfer is in agreement with EU legislation on the general data protection regulation and decisions by the Ethical Review Board of Sweden and Region Skåne, which should be regulated in a material transfer agreement.

## Author contributions

JLR, GS, LMG, HZ, KB and OH created the concept and design. Data acquisition was performed by JLR, GS, LMG, AOD, LSBW, JV, ES and SP. JLR, LMG and GS performed data analysis. JLR, GS, LMG, SJ, HZ, KB and OH contributed to sample selection. All authors contributed to the interpretation of data. JLR, LMG and GS drafted the manuscript, and all authors revised. All authors read and approved the final manuscript.

## Data Availability

Anonymized data will be shared by request from a qualified academic investigator for the sole purpose of replicating procedures and results presented in the article and as long as data transfer is in agreement with EU legislation on the general data protection regulation and decisions by the Ethical Review Board of Sweden and Region Skane, which should be regulated in a material transfer agreement.

## Acknowledgments

The authors would like to express their sincere gratitude to the BioFINDER-1 and BioFINDER-2 participants and relatives, without whom this research would have not been possible. The authors thank the various foundations that kindly supported this research.

## REFERENCES

1. Dubois, B., et al., Preclinical Alzheimer’s disease: Definition, natural history, and diagnostic criteria. Alzheimers Dement, 2016. 12(3): p. 292–323.

2. Scheltens, P., et al., Alzheimer’s disease. Lancet, 2016. 388(10043): p. 505–17.

3. Blennow, K., M.J. de Leon, and H. Zetterberg, Alzheimer’s disease. Lancet, 2006. 368(9533): p. 387–403.

4. Masters, C.L., et al., Alzheimer’s disease. Nat Rev Dis Primers, 2015. 1: p. 15056.

5. Hansson, O., Biomarkers for neurodegenerative diseases. Nat Med, 2021. 27(6): p. 954–963.

6. Palmqvist, S., et al., Cerebrospinal fluid analysis detects cerebral amyloid-beta accumulation earlier than positron emission tomography. Brain, 2016. 139(Pt 4): p. 1226–36.

7. Gordon, B.A., et al., Spatial patterns of neuroimaging biomarker change in individuals from families with autosomal dominant Alzheimer’s disease: a longitudinal study. Lancet Neurol, 2018. 17(3): p. 241–250.

8. Villemagne, V.L., et al., Amyloid beta deposition, neurodegeneration, and cognitive decline in sporadic Alzheimer’s disease: a prospective cohort study. Lancet Neurol, 2013. 12(4): p. 357–67.

9. Scholl, M., et al., Biomarkers for tau pathology. Mol Cell Neurosci, 2019. 97: p. 18–33.

10. Blennow, K. and H. Zetterberg, Biomarkers for Alzheimer’s disease: current status and prospects for the future. J Intern Med, 2018. 284(6): p. 643–663.

11. Wolk, D.A., et al., Use of Flutemetamol F 18-Labeled Positron Emission Tomography and Other Biomarkers to Assess Risk of Clinical Progression in Patients With Amnestic Mild Cognitive Impairment. JAMA Neurol, 2018. 75(9): p. 1114–1123.

12. Rabinovici, G.D., et al., Association of Amyloid Positron Emission Tomography With Subsequent Change in Clinical Management Among Medicare Beneficiaries With Mild Cognitive Impairment or Dementia. JAMA, 2019. 321(13): p. 1286–1294.

13. Lantero-Rodriguez, J., et al., CSF p-tau205: a biomarker of tau pathology in Alzheimer’s disease. Acta Neuropathol, 2024. 147(1): p. 12.

14. Lantero-Rodriguez, J., et al., Biofluid-based staging of Alzheimer’s disease. Acta Neuropathol, 2025. 149(1): p. 27.

15. Montoliu-Gaya, L., et al., Optimal blood tau species for the detection of Alzheimer’s disease neuropathology: an immunoprecipitation mass spectrometry and autopsy study. Acta Neuropathol, 2023. 147(1): p. 5.

16. Montoliu-Gaya, L., et al., Mass spectrometric simultaneous quantification of tau species in plasma shows differential associations with amyloid and tau pathologies. Nat Aging, 2023. 3(6): p. 661–669.

17. Barthelemy, N.R., et al., CSF tau phosphorylation occupancies at T217 and T205 represent improved biomarkers of amyloid and tau pathology in Alzheimer’s disease. Nat Aging, 2023. 3(4): p. 391–401.

18. Horie, K., et al., Plasma MTBR-tau243 biomarker identifies tau tangle pathology in Alzheimer’s disease. Nat Med, 2025.

19. Braak, H., et al., Staging of Alzheimer disease-associated neurofibrillary pathology using paraffin sections and immunocytochemistry. Acta Neuropathol, 2006. 112(4): p. 389–404.

20. Gandhi, N.S., et al., A Phosphorylation-Induced Turn Defines the Alzheimer’s Disease AT8 Antibody Epitope on the Tau Protein. Angew Chem Int Ed Engl, 2015. 54(23): p. 6819–23.

21. Malia, T.J., et al., Epitope mapping and structural basis for the recognition of phosphorylated tau by the anti-tau antibody AT8. Proteins, 2016. 84(4): p. 427–34.

22. Goedert, M., R. Jakes, and E. Vanmechelen, Monoclonal antibody AT8 recognises tau protein phosphorylated at both serine 202 and threonine 205. Neurosci Lett, 1995. 189(3): p. 167–9.

23. Gobom, J., et al., Antibody-free measurement of cerebrospinal fluid tau phosphorylation across the Alzheimer’s disease continuum. Mol Neurodegener, 2022. 17(1): p. 81.

24. Barthelemy, N.R., et al., Tau Phosphorylation Rates Measured by Mass Spectrometry Differ in the Intracellular Brain vs. Extracellular Cerebrospinal Fluid Compartments and Are Differentially Affected by Alzheimer’s Disease. Front Aging Neurosci, 2019. 11: p. 121.

25. Barthelemy, N.R., et al., Site-Specific Cerebrospinal Fluid Tau Hyperphosphorylation in Response to Alzheimer’s Disease Brain Pathology: Not All Tau Phospho-Sites are Hyperphosphorylated. J Alzheimers Dis, 2022. 85(1): p. 415–429.

26. Barthelemy, N.R., et al., A soluble phosphorylated tau signature links tau, amyloid and the evolution of stages of dominantly inherited Alzheimer’s disease. Nat Med, 2020. 26(3): p. 398–407.

27. Salvado, G., et al., Disease staging of Alzheimer’s disease using a CSF-based biomarker model. Nat Aging, 2024. 4(5): p. 694–708.

28. Therriault, J., et al., Biomarker-based staging of Alzheimer disease: rationale and clinical applications. Nat Rev Neurol, 2024. 20(4): p. 232–244.

29. Jack, C.R., Jr., et al., Revised criteria for diagnosis and staging of Alzheimer’s disease: Alzheimer’s Association Workgroup. Alzheimers Dement, 2024. 20(8): p. 5143–5169.

30. Petrazzuoli, F., et al., Brief Cognitive Tests Used in Primary Care Cannot Accurately Differentiate Mild Cognitive Impairment from Subjective Cognitive Decline. J Alzheimers Dis, 2020. 75(4): p. 1191–1201.

31. Palmqvist, S., et al., Cognitive effects of Lewy body pathology in clinically unimpaired individuals. Nat Med, 2023. 29(8): p. 1971–1978.

32. Dubois, B., et al., Clinical diagnosis of Alzheimer’s disease: recommendations of the International Working Group. Lancet Neurol, 2021. 20(6): p. 484–496.

33. Palmqvist, S., et al., Discriminative Accuracy of Plasma Phospho-tau217 for Alzheimer Disease vs Other Neurodegenerative Disorders. JAMA, 2020. 324(8): p. 772–781.

34. Janelidze, S., et al., Head-to-head comparison of 10 plasma phospho-tau assays in prodromal Alzheimer’s disease. Brain, 2023. 146(4): p. 1592–1601.

35. Ashton, N.J., et al., Differential roles of Abeta42/40, p-tau231 and p-tau217 for Alzheimer’s trial selection and disease monitoring. Nat Med, 2022. 28(12): p. 2555–2562.

36. Ossenkoppele, R., et al., Amyloid and tau PET-positive cognitively unimpaired individuals are at high risk for future cognitive decline. Nat Med, 2022. 28(11): p. 2381–2387.

37. Jack, C.R., Jr., et al., Different definitions of neurodegeneration produce similar amyloid/neurodegeneration biomarker group findings. Brain, 2015. 138(Pt 12): p. 3747–59.

38. Ikari, Y., et al., Improved Correlation of (18)F-Flortaucipir PET SUVRs and Clinical Stages in the Alzheimer Disease Continuum with the MUBADA/PERSI-Based Analysis. J Nucl Med Technol, 2024. 52(4): p. 340–347.

39. Moloney, C.M., V.J. Lowe, and M.E. Murray, Visualization of neurofibrillary tangle maturity in Alzheimer’s disease: A clinicopathologic perspective for biomarker research. Alzheimers Dement, 2021. 17(9): p. 1554–1574.

40. Pontecorvo, M.J., et al., Association of Donanemab Treatment With Exploratory Plasma Biomarkers in Early Symptomatic Alzheimer Disease: A Secondary Analysis of the TRAILBLAZER-ALZ Randomized Clinical Trial. JAMA Neurol, 2022. 79(12): p. 1250–1259.

