## Supplementary Material for "P-TAU205 IS A BIOMARKER LINKED TO TAU-PET ABNORMALITY: A CROSS-SECTIONAL AND LONGITUDINAL STUDY"

**Supplementary Figure 1: In BioFINDER-1, CSF p-tau205 concentrations in by clinical diagnosis (A) and cross-sectional associations between CSF p-tau205 with Aβ-PET (B), cortical thickness (C) and MMSE (D).** Differences in CSF p-tau205 levels by groups were determined using ANCOVA and Tukey's method for post-hoc comparisons. Age and sex (and years of education for cognitive outcomes) were used as covariates in all cases. Box plots include all participants, displaying the median and the interquartile range; whiskers show the lower value of maximum/minimum value or 1.5 interquartile range from the hinge. Linear regressions with CSF p-tau205 as predictor and PET, MRI and MMSE as outcome were used to measure the association. Non-AD individuals were excluded from analyses with cortical thickness or cognition. Standardized β (βstd) and p-values of the associations as well as the R2 of the model for the Aβ-positive individuals are shown in Supplementary Table 5. *: $p < 0.05$; **: $p < 0.01$; ***: $p < 0.001$.

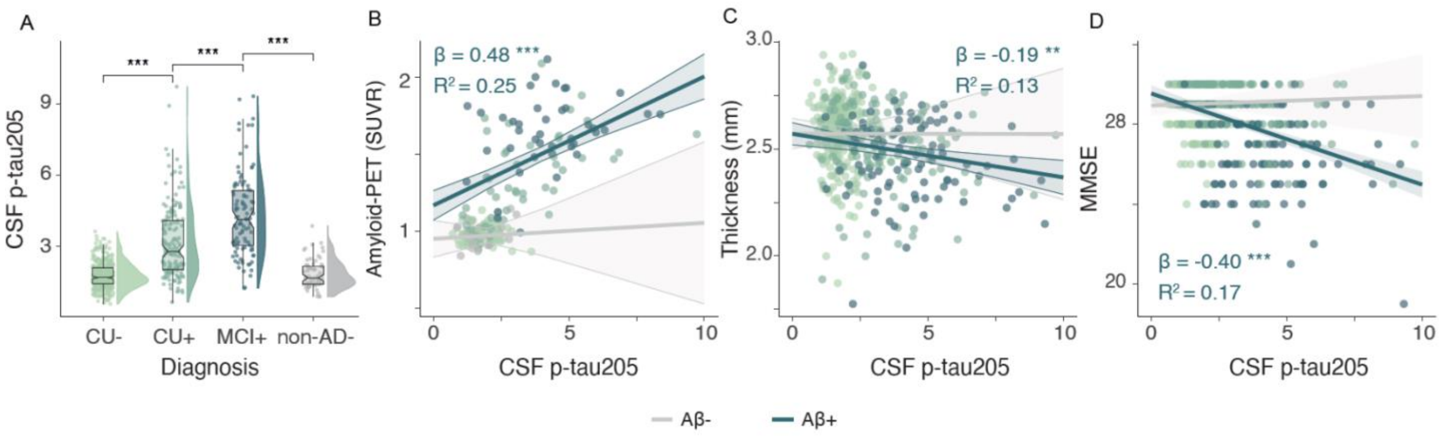

**Supplementary Table 1: CSF p-tau205 differences by diagnosis in BioFINDER-2.**
Fold changes (p-values) are shown in the table. ANCOVA were used to assess statistical differences adjusting for age and sex.

| | CU- | CU+ | MCI+ | ADdem | nonAD+ |
|---|---|---|---|---|---|
| CU- | - | - | - | - | - |
| CU+ | 1.91 (<0.001) | - | - | - | - |
| MCI+ | 2.32 (<0.001) | 1.21 (<0.001) | - | - | - |
| ADdem | 2.95 (<0.001) | 1.54 (<0.001) | 1.27 (<0.001) | - | - |
| nonAD+ | 1.65 (<0.001) | 0.87 (0.108) | 0.71 (<0.001) | 0.56 (<0.001) | - |
| nonAD- | 1.1 (0.835) | 0.58 (<0.001) | 0.47 (<0.001) | 0.37 (<0.001) | 0.66 (<0.001) |

**Supplementary Table 2: CSF p-tau205 differences by A/T status in BioFINDER-2.**
Fold changes (p-values) are shown in the table. ANCOVA were used to assess statistical differences adjusting for age and sex.

| | A-T- | A+T- | A+T+ |
|---|---|---|---|
| A-T- | | | |
| A+T- | 1.7 (<0.001) | | |
| A+T+ | 2.76 (<0.001) | 1.62 (<0.001) | |
| A-T+ | 1.15 (0.916) | 0.67 (0.123) | 0.42 (<0.001) |

**Supplementary Table 3: CSF p-tau205 differences by Braak stages in BioFINDER-2.**
Fold changes (p-values) are shown in the table. ANCOVA were used to assess statistical differences adjusting for age and sex.

| | 0 | I-II | III-IV | V-VI |
|---|---|---|---|---|
| 0 | | | | |
| I-II | 1.68 (<0.001) | | | |
| III-IV | 2.26 (<0.001) | 1.35 (<0.001) | | |
| V-VI | 2.62 (<0.001) | 1.56 (<0.001) | 1.16 (<0.001) | |

**Supplementary Table 4: CSF p-tau205 differences by biological categorization using the AA criteria in BioFINDER-2.**
Fold changes (p-values) are shown in the table. ANCOVA were used to assess statistical differences adjusting for age and sex.

| | A- | A+ | AMTL+ | AMTLN+ |
|---|---|---|---|---|
| A- | | | | |
| A+ | 1.55 (<0.001) | | | |
| AMTL+ | 2.17 (<0.001) | 1.4 (<0.001) | | |
| AMTLN+ | 2.54 (<0.001) | 1.64 (<0.001) | 1.17 (0.012) | |
| AMTLN++ | 2.97 (<0.001) | 1.92 (<0.001) | 1.37 (<0.001) | 1.17 (<0.001) |

**Supplementary Table 5: Cross-sectional associations between CSF p-tau205 and AD biomarkers by Aβ-status.**
Linear regression models were used adjusting for age and sex (and years of education for cognitive outcomes). Independent models were created for Aβ-negative and Aβ-positive individuals. Non-AD individuals were excluded from analyses with cortical thickness or cognition as outcomes.

| | Aβ-negative | | | | Aβ-positive | | | |
|---|---|---|---|---|---|---|---|---|
| | N | β[95%CI] | p-value | $R^2$ | N | β[95%CI] | p-value | $R^2$ |
| BioFINDER-2 | | | | | | | | |
| Aβ-PET | 558 | 0.06 [-0.03, 0.14] | 0.18 | 0.05 | 353 | 0.49 [0.4, 0.58] | <0.001 | 0.26 |
| MTL Tau-PET | 724 | 0.18 [0.11, 0.25] | 0 | 0.07 | 606 | 0.56 [0.5, 0.63] | <0.001 | 0.32 |
| NeoT Tau-PET | 725 | 0.08 [0.01, 0.15] | 0.031 | 0.06 | 607 | 0.5 [0.43, 0.57] | <0.001 | 0.29 |
| Cortical thickness | 447 | 0.01 [-0.08, 0.1] | 0.828 | 0.17 | 530 | -0.29 [-0.37, -0.21] | <0.001 | 0.14 |
| mPACC | 454 | 0.04 [-0.04, 0.12] | 0.339 | 0.31 | 496 | -0.33 [-0.41, -0.24] | <0.001 | 0.14 |
| MMSE | 457 | 0 [-0.1, 0.09] | 0.941 | 0.05 | 534 | -0.34 [-0.42, -0.26] | <0.001 | 0.14 |
| BioFINDER-1 | | | | | | | | |
| Aβ-PET | 102 | 0.08 [-0.11, 0.28] | 0.406 | 0.01 | 86 | 0.48 [0.29, 0.67] | <0.001 | 0.25 |
| Cortical thickness | 255 | 0 [-0.12, 0.11] | 0.95 | 0.11 | 222 | -0.19 [-0.31, -0.06] | 0.003 | 0.13 |
| mPACC | 268 | 0.08 [-0.03, 0.2] | 0.159 | 0.08 | 223 | -0.43 [-0.55, -0.31] | <0.001 | 0.21 |
| MMSE | 379 | 0.03 [-0.07, 0.13] | 0.567 | 0.03 | 261 | -0.4 [-0.52, -0.29] | <0.001 | 0.17 |

**Supplementary Table 6: Longitudinal associations between baseline CSF p-tau205 and AD biomarker changes.**
Linear mixed models were used with random intercepts and time slopes, adjusting for age and sex (and years of education for cognitive outcomes). Non-AD individuals were excluded from analyses with cortical thickness or cognition as outcomes.

| | N | β[95%CI] | p-value | $R^2$ |
|---|---|---|---|---|
| BioFINDER-2 | | | | |
| Aβ-PET | 395 | 0.04 [0.03, 0.05] | <0.001 | 0.44 |
| MTL Tau-PET | 495 | 0.05 [0.04, 0.06] | <0.001 | 0.50 |
| NeoT Tau-PET | 495 | 0.12 [0.1, 0.14] | <0.001 | 0.33 |
| BioFINDER-1 | | | | |
| Cortical thickness | 450 | -0.17 [-0.19, -0.14] | <0.001 | 0.24 |
| mPACC | 468 | -0.32 [-0.36, -0.28] | <0.001 | 0.38 |
| MMSE | 650 | -0.4 [-0.45, -0.36] | <0.001 | 0.36 |

**Supplementary Table 7: Longitudinal CSF p-tau205 changes by Aβ-status.**
Linear mixed models were used with random intercepts and time slopes, adjusting for age and sex.

| | N | β[95%CI] | p-value | $R^2$ |
|---|---|---|---|---|
| All | 714 | 0.14 [0.11, 0.18] | <0.001 | 0.44 |
| CU | 544 | 0.16 [0.12, 0.21] | <0.001 | 0.35 |
| CI | 170 | 0.10 [0.04, 0.16] | <0.001 | 0.57 |

**Supplementary Table 8: Associations between longitudinal CSF p-tau205 changes and longitudinal AD biomarker changes in BioFINDER-1.**
Longitudinal CSF p-tau205 changes and longitudinal changes of AD biomarkers were calculated by linear mixed models, independently, with random intercepts and time slopes. Linear regression models were used to assess associations between longitudinal CSF p-tau205 changes and AD biomarker changes, adjusting for age and sex (and years of education for cognitive outcomes). Non-AD individuals were excluded from these analyses.

| | N | β[95%CI] | p-value | $R^2$ |
|---|---|---|---|---|
| Cortical thickness | 698 | -0.47 [-0.54, -0.4] | <0.001 | 0.32 |
| MMSE | 698 | -0.6 [-0.66, -0.54] | <0.001 | 0.41 |
| mPACC | 698 | -0.61 [-0.68, -0.55] | <0.001 | 0.46 |

**Supplementary Table 9: Cross-sectional associations between baseline CSF stages and AD biomarker changes.**

Linear regression models were used adjusting for age and sex (and years of education for cognitive outcomes). We show comparisons against contiguous CSF stages. Non-AD individuals were excluded from analyses with cortical thickness or cognition as outcomes.

| | β[95%CI] stage 1vs 0 | p-value | β[95%CI] stage 2 vs 1 | p-value | β[95%CI] stage 3 vs 2 | p-value |
|---|---|---|---|---|---|---|
| BioFINDER-2 | | | | | | |
| Aβ-PET | 0.6 [0.43, 0.78] | <0.001 | 0.87 [0.63, 1.11] | <0.001 | 0.45 [0.25, 0.64] | <0.001 |
| MTL Tau-PET | 0.11 [-0.08, 0.3] | 0.425 | 0.7 [0.45, 0.95] | <0.001 | 0.79 [0.59, 0.99] | <0.001 |
| NeoT Tau-PET | 0.08 [-0.15, 0.31] | 0.794 | 0.44 [0.14, 0.74] | 0.001 | 0.81 [0.58, 1.05] | <0.001 |
| Cortical thickness | -0.19 [-0.47, 0.08] | 0.268 | -0.28 [-0.62, 0.06] | 0.152 | -0.41 [-0.67, -0.16] | <0.001 |
| mPACC | -0.24 [-0.5, 0.02] | 0.075 | -0.62 [-0.94, -0.29] | <0.001 | -0.32 [-0.57, -0.08] | 0.005 |
| MMSE | -0.15 [-0.44, 0.14] | 0.537 | -0.58 [-0.94, -0.22] | <0.001 | -0.35 [-0.62, -0.07] | 0.006 |
| BioFINDER-1 | | | | | | |
| Cortical thickness | -0.22 [-0.6, 0.16] | 0.436 | 0.03 [-0.5, 0.55] | 0.999 | -0.39 [-0.83, 0.05] | 0.101 |
| mPACC | -0.22 [-0.55, 0.1] | 0.276 | -0.51 [-0.98, -0.05] | 0.022 | -0.52 [-0.92, -0.13] | 0.004 |
| MMSE | -0.03 [-0.35, 0.3] | 0.997 | -0.47 [-0.93, -0.02] | 0.038 | -0.5 [-0.89, -0.11] | 0.005 |

**Supplementary Table 10: Longitudinal associations between baseline CSF stages and AD biomarker changes.**
Linear mixed models were used with random intercepts and time slopes, adjusting for age and sex (and years of education for cognitive outcomes). We show comparisons against CSF stage 0 in all cases. Non-AD individuals were excluded from analyses with cortical thickness or cognition as outcomes.

| | N | β[95%CI] stage 1 | p-value | β[95%CI] stage 2 | p-value | β[95%CI] stage 3 | p-value | $R^2$ |
|---|---|---|---|---|---|---|---|---|
| BioFINDER-2 | | | | | | | | |
| Aβ-PET | 420 | 0.12 [0.09, 0.15] | <0.001 | 0.15 [0.1, 0.19] | <0.001 | 0.11 [0.09, 0.14] | <0.001 | 0.73 |
| MTL Tau-PET | 598 | 0.06 [0.03, 0.1] | <0.001 | 0.14 [0.1, 0.18] | <0.001 | 0.13 [0.11, 0.15] | <0.001 | 0.56 |
| NeoT Tau-PET | 598 | 0.04 [-0.01, 0.09] | 0.138 | 0.13 [0.07, 0.19] | <0.001 | 0.27 [0.23, 0.3] | <0.001 | 0.35 |
| Cortical thickness | 467 | -0.06 [-0.12, 0.01] | 0.087 | -0.21 [-0.28, -0.13] | <0.001 | -0.26 [-0.3, -0.22] | <0.001 | 0.37 |
| mPACC | 520 | -0.03 [-0.15, 0.08] | 0.587 | -0.17 [-0.31, -0.04] | 0.013 | -0.44 [-0.52, -0.36] | <0.001 | 0.44 |
| MMSE | 520 | -0.06 [-0.21, 0.09] | 0.437 | -0.11 [-0.29, 0.06] | 0.211 | -0.64 [-0.74, -0.53] | <0.001 | 0.36 |
| BioFINDER-1 | | | | | | | | |
| Cortical thickness | 428 | -0.06 [-0.14, 0.01] | 0.098 | -0.11 [-0.2, -0.02] | 0.014 | -0.38 [-0.43, -0.33] | <0.001 | 0.26 |
| mPACC | 442 | -0.13 [-0.24, -0.01] | 0.032 | -0.25 [-0.39, -0.11] | <0.001 | -0.83 [-0.91, -0.74] | <0.001 | 0.42 |
| MMSE | 608 | -0.09 [-0.23, 0.05] | 0.22 | -0.39 [-0.55, -0.23] | <0.001 | -0.95 [-1.05, -0.86] | <0.001 | 0.37 |

**Supplementary Table 11: Progression to dementia by baseline CSF stages**
Proportional Hazard-Cox models were used adjusting for age, sex and baseline clinical stage (cognitively unimpaired or MCI).

| CSF stages | HR[95%CI] | p-value |
|---|---|---|
| Stage 1 | 1.89 [0.90, 3.93] | 0.091 |
| Stage 2 | 3.18 [1.70, 5.94] | <0.001 |
| Stage 3 | 6.40 [4.28, 9.59] | <0.001 |
